# Reduced COVID-19 Hospitalizations among New York City Residents Following Age-Based SARS-CoV-2 Vaccine Eligibility: Evidence from a Regression Discontinuity Design

**DOI:** 10.1101/2021.06.30.21259491

**Authors:** Sharon K. Greene, Alison Levin-Rector, Emily McGibbon, Jennifer Baumgartner, Katelynn Devinney, Alexandra Ternier, Jessica Sell, Rebecca Kahn, Nishant Kishore

## Abstract

**Background:** In clinical trials, several SARS-CoV-2 vaccines were shown to reduce risk of severe COVID-19 illness. Local, population-level, real-world evidence of vaccine effectiveness is accumulating. We assessed vaccine effectiveness for community-dwelling New York City (NYC) residents using a quasi-experimental, regression discontinuity design, leveraging a period (January 12–March 9, 2021) when ≥65-year-olds were vaccine-eligible but younger persons, excluding essential workers, were not.

**Methods:** We constructed segmented, negative binomial regression models of age-specific COVID-19 hospitalization rates among 45–84-year-old NYC residents during a post-vaccination program implementation period (February 21–April 17, 2021), with a discontinuity at age 65 years. The relationship between age and hospitalization rates in an unvaccinated population was incorporated using a pre-implementation period (December 20, 2020–February 13, 2021). We calculated the rate ratio (RR) and 95% confidence interval (CI) for the interaction between implementation period (pre or post) and age-based eligibility (45–64 or 65–84 years). Analyses were stratified by race/ethnicity and borough of residence. Similar analyses were conducted for COVID-19 deaths.

**Results:** Hospitalization rates among 65–84-year-olds decreased from pre- to post-implementation periods (RR 0.85, 95% CI: 0.74–0.97), controlling for trends among 45–64-year-olds. Accordingly, an estimated 721 (95% CI: 126–1,241) hospitalizations were averted. Residents just above the eligibility threshold (65–66-year-olds) had lower hospitalization rates than those below (63–64-year-olds). Racial/ethnic groups and boroughs with higher vaccine coverage generally experienced greater reductions in RR point estimates. Uncertainty was greater for the decrease in COVID-19 death rates (RR 0.85, 95% CI: 0.66–1.10).

**Conclusion:** The vaccination program in NYC reduced COVID-19 hospitalizations among the initially age-eligible ≥65-year-old population by approximately 15%. The real-world evidence of vaccine effectiveness makes it more imperative to improve vaccine access and uptake to reduce inequities in COVID-19 outcomes.

## 1. Introduction

The SARS-CoV-2 vaccines authorized and recommended for emergency use in the United States were demonstrated in randomized clinical trials to reduce risk of severe COVID-19 illness [1-3]. Post-authorization, several studies have demonstrated real-world evidence of SARS-CoV-2 vaccine effectiveness in various settings, such as among skilled nursing facility residents and essential workers [4, 5], or at the national level [6, 7]. Local, population-level evidence of effectiveness can support public messaging to promote the importance of vaccination [8].

In New York City (NYC), guidelines for vaccine eligibility were established by the Office of the Governor of New York State. When vaccinations began on December 14, 2020 [9], eligibility was initially restricted to health care workers and residents and staff of long-term care facilities. Eligibility expanded to ≥75-year-olds in the general population and essential workers (workers in education, public safety, and public transit, and first responders) on January 11, 2021 [10], to ≥65-year-olds on January 12 [11], to ≥60-year-olds and additional categories of public-facing essential workers on March 10 [12], to ≥50-year-olds and individuals with comorbidities and underlying conditions on March 23 [13], to ≥30-year-olds on March 30 [14], to ≥16-year-olds on April 6 [14], and to ≥12-year-olds on May 12 [15]. Screeners at vaccination sites verified age-based eligibility by requiring proof of age, such as a driver’s license, IDNYC (a free municipal identification card for NYC residents), birth certificate, passport, permanent resident card, certificate of naturalization or citizenship, or life insurance policy or marriage certificate with birthdate [16]. Notably, vaccine eligibility for ≥65-year-olds as of mid-January coincided with the second peak of COVID-19 hospitalizations in NYC [17], such that the vaccination program was established concurrently with a waning epidemic period.

Vaccinees and non-vaccinees are likely systematically different in ways that are difficult to observe (e.g., adherence to social distancing recommendations and presence of underlying conditions) yet influence their probability of SARS-CoV-2 infection and testing and COVID-19 hospitalization and death [18]. Cross-sectional studies comparing outcome rates among vaccinated and unvaccinated populations are susceptible to confounding due to population differences arising from volunteer selection bias, healthy vaccinee effects, and frailty bias [19].

The NYC Department of Health and Mental Hygiene (DOHMH) sought to assess evidence of population-level vaccine effectiveness in NYC, citywide and stratified by subpopulations with different vaccination coverage. We used a quasi-experimental observational study design to leverage an 8-week period (January 12–March 9, 2021) when community residents at an age threshold of ≥65-years were vaccine-eligible but younger persons (excluding essential workers) were not.

## 2. Methods

### 2.1 COVID-19 hospitalization and death data

Confirmed and probable cases of COVID-19 [20] among NYC residents are reported to NYC DOHMH through electronic laboratory reporting, and hospitalizations and deaths for these patients are ascertained by routinely importing and matching data from supplemental systems, as previously described [21]. COVID-19 hospitalizations were defined as NYC residents admitted within +/- 14 days of the first date of specimen collection that tested positive for SARS-CoV-2 by a molecular or antigen test; or if not laboratory-positive but a symptomatic contact of a confirmed or probable case, then admitted within +/- 14 days of illness onset. Hospitalizations with missing admission date (n = 940, 3.3%) were omitted from analysis. COVID-19 deaths were defined as NYC residents who had a positive molecular test and (a) the cause-of-death on the death certificate was COVID-19 or similar, or b) COVID-19 was not a cause-of-death on the death certificate but the patient died within 60 days of a positive molecular test, and the death was not due to external causes such as injury (“confirmed deaths”); or the cause-of-death on the death certificate was COVID-19 or similar, but a positive molecular test was not reported (“probable deaths”) [21]. Patient age was calculated as of January 12, 2021, not as of hospitalization or death date.

Two categories of patients were excluded from analysis. First, given low hospitalization and death rates [22], patients <45 years-old as of January 12, 2021 were excluded from the comparator for trends among vaccine-eligible ≥65-year-olds. Patients ≥85 years-old were also excluded for sparsity, such that the study population was restricted to 45–84-year-olds, i.e., +/- 20 years around the vaccine eligibility threshold of age 65 years. Second, patients residing in congregate settings (e.g., long-term care or correctional facilities) were excluded because their vaccine eligibility timing was different from community residents and less dependent on age. Such patients were identified by geocoding the residential address at time of report and matching to facility lists.

### 2.2 Vaccination and population denominator data

The cumulative percentage of NYC residents having received at least the first dose of a SARS-CoV-2 vaccine, by vaccination date and age at first dose, was obtained from the NYC DOHMH Citywide Immunization Registry [23], as reported by immunizing facilities. Single-year of age population estimates for 2019 for the five NYC boroughs (equivalent to counties) were downloaded from the National Center for Health Statistics [24].

### 2.3 Program implementation timing

We assumed that population-level vaccine effects on hospitalizations would not be apparent until 4 weeks after age-based eligibility was established. Within that period, we accounted for 1 week to begin substantial vaccination uptake in the newly eligible population, an additional 2 weeks after receipt of the first dose for a partially protective effect from vaccination, and an additional 1 week for hospitalizations to occur among those infected. Following the same logic, we assumed that vaccine effects on deaths would not be apparent until 6 weeks after age-based eligibility was established, additionally accounting for an average lag of approximately 2 weeks between COVID-19 hospitalization and death.

We defined pre- and post-vaccination program implementation periods of 8 weeks each, defining weeks as Sundays–Saturdays. We chose 8 weeks to correspond with the duration of the period (January 12–March 9, 2021) when only ≥65-year-olds had age-based eligibility. For the primary analysis for hospitalizations, we defined the pre-implementation period as December 20, 2020–February 13, 2021, i.e., an 8-week period ending 4 weeks after ≥65-year-olds became eligible on Jan. 12. Imposing a 1-week washout period, we defined the post-implementation period as February 21–April 17, 2021. For the primary analysis for deaths, we defined the pre-implementation period as January 3–February 27, 2021, i.e., an 8-week period ending 6 weeks after ≥65-year-olds became eligible on Jan. 12. Imposing a 2-week washout period to account for additional ambiguity in the timing of vaccine effects, we defined the post-implementation period as March 14–May 8, 2021. Data were frozen as of June 28, 2021, capturing hospitalizations ascertained within 72 days and deaths ascertained within 51 days after the ends of the post-implementation periods for the primary analysis. In sensitivity analyses, we shifted implementation period definitions and imposed washout periods of different lengths.

### 2.4 Regression discontinuity design

We constructed segmented, negative binomial regression models of the age-specific hospitalization (and death) rates during the post-vaccination program implementation period, with a discontinuity at age 65 years. We used pre-implementation period data to incorporate the observed relationship between age and hospitalization rates in an unvaccinated population. We expected the overall hospitalization rate in the post-implementation period to be lower due to the waning stage of the epidemic, but for trends across age to persist. We specified the model as a standard regression discontinuity design with a control group and indexed and centered the values for age and their corresponding interaction terms for appropriate interpretations of regression coefficients of interest (Appendix A) [25]. The analytic dataset for the hospitalizations primary analysis is provided for reproducibility (Appendix B). Analyses were conducted using PROC GENMOD in SAS Version 9.4 (SAS Institute, Cary, NC). This work was deemed public health surveillance that is non-research by the NYC DOHMH Institutional Review Board.

The key parameter of interest was *β*_*6*_, the interaction term between vaccine program implementation period (pre or post) and age-based eligibility (45–64 or 65–84 years), representing the adjusted difference in log rates (intercept change) for 65-year-olds following age-based eligibility [26-29] (Appendix A). We exponentiated this parameter estimate and 95% confidence interval (CI) to obtain the rate ratio (RR) of interest and 95% CI.

### 2.5 Estimating hospitalizations and deaths averted

We estimated hospitalizations and deaths among 65–84-year-olds during the post-implementation period under the counterfactual scenario in which there were no effects of vaccination. That is, we estimated hospitalizations and deaths in this group had they experienced the same intercept and slope change from the pre- to post-implementation periods as the 45–64-year-olds but experienced no discontinuity due to the implementation of the age-based vaccination policy. As above, let *β*_6_ equal the parameter of a fitted negative binomial model describing the log difference of hospitalization rates between pre- and post-implementation periods among 65–84-year-olds, controlling for the decrease in hospitalizations in the post-implementation period due to the waning epidemic. Therefore, *e*^β6^ is the rate ratio of this parameter.

Let *Y*_65–84_ equal the observed hospitalization rate among 65–84-year-olds in the post-implementation period and *N*_65–84_ equal the total population of 65–84-year-olds in NYC. Therefore, the counterfactual scenario in which there was no change in the hospitalization rate between 65–84-year-olds in the pre- and post-implementation periods, controlling for the decrease in hospitalizations in the post-implementation period is:

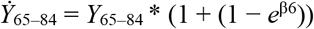

and the expected number of hospitalizations in the counterfactual is:

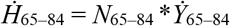

while the observed number of hospitalizations is:

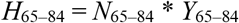

Finally, assuming *β*_6_ is negative, the number of averted hospitalizations is defined as:

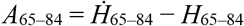

Therefore, the final equation of averted hospitalizations is:

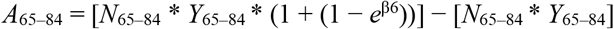

Similarly, for the 95% CI, we replaced *β*_6_ with the upper and lower limits of the parameter’s CI. The same approach was used to calculate averted deaths.

### 2.6 Stratified analyses and negative controls

To assess heterogeneity of findings for hospitalizations across subpopulations with different vaccination rates, stratified analyses were conducted by race/ethnicity and borough of residence, where non-missing. Stratified analyses were not conducted for deaths given sparsity, nor for sub-borough areas because borough was the smallest geographic resolution available with population denominators by single year of age [24].

We used negative controls to assess whether findings for citywide COVID-19 hospitalizations might be attributable to unknown sources of error [30]. If the vaccination program were effective, then these methods should demonstrate reduced COVID-19 hospitalization rates when applied at the ≥65 years age threshold during the period around vaccine program implementation, but null effects when applied to a different age threshold or to earlier periods when no SARS-CoV-2 vaccines were available.

First, we modified our primary analysis by redefining the age groups from 45–64 and 65– 84 (corresponding to the true age threshold of ≥65 years) to 30–49 and 50–79 (corresponding to a false age threshold of ≥50 years). Second, we modified the primary analysis by shifting the 16-week study period with 1-week washout period to four negative control points earlier in the epidemic based on trends in citywide hospitalizations [17], defining the start of the post-implementation period as May 10, 2020 (when hospitalizations were at a similar magnitude and waning during the first epidemic wave), August 16, 2020 (when hospitalizations were low between epidemic waves), December 20, 2020 (when hospitalizations were waxing during the second wave), and January 24, 2021 (when hospitalizations were steady during the second wave prior to widespread community vaccine availability).

## 3. Results

### 3.1 Trends in vaccination coverage and COVID-19 hospitalizations

As of January 16, 2021 (the end of the week age-based eligibility began for ≥65-year-olds), the cumulative percentage of 65–84-year-old NYC residents having received at least the first dose of a SARS-CoV-2 vaccine was 6.5% (Figure 1). By March 9 (the last day prior to ≥60-year-olds also becoming vaccine-eligible), 47.6% of 65–84-year-olds were vaccinated, compared with only 22.2% of 45–64-year-olds. Vaccination coverage of 65–84-year-olds by March 9 varied widely by race/ethnicity, ranging from 26.2% of Black/African-American NYC residents to 45.9% of Asian/Pacific Islander NYC residents. Vaccination coverage of 65–84-year-olds as of March 20 (4 weeks before the end of the post-implementation period) was 55.8%. During the second COVID-19 wave in NYC, the timing of peak hospitalizations was similar for younger (45–64) and older (65–84) age groups (Figure 1).

**Figure 1.**
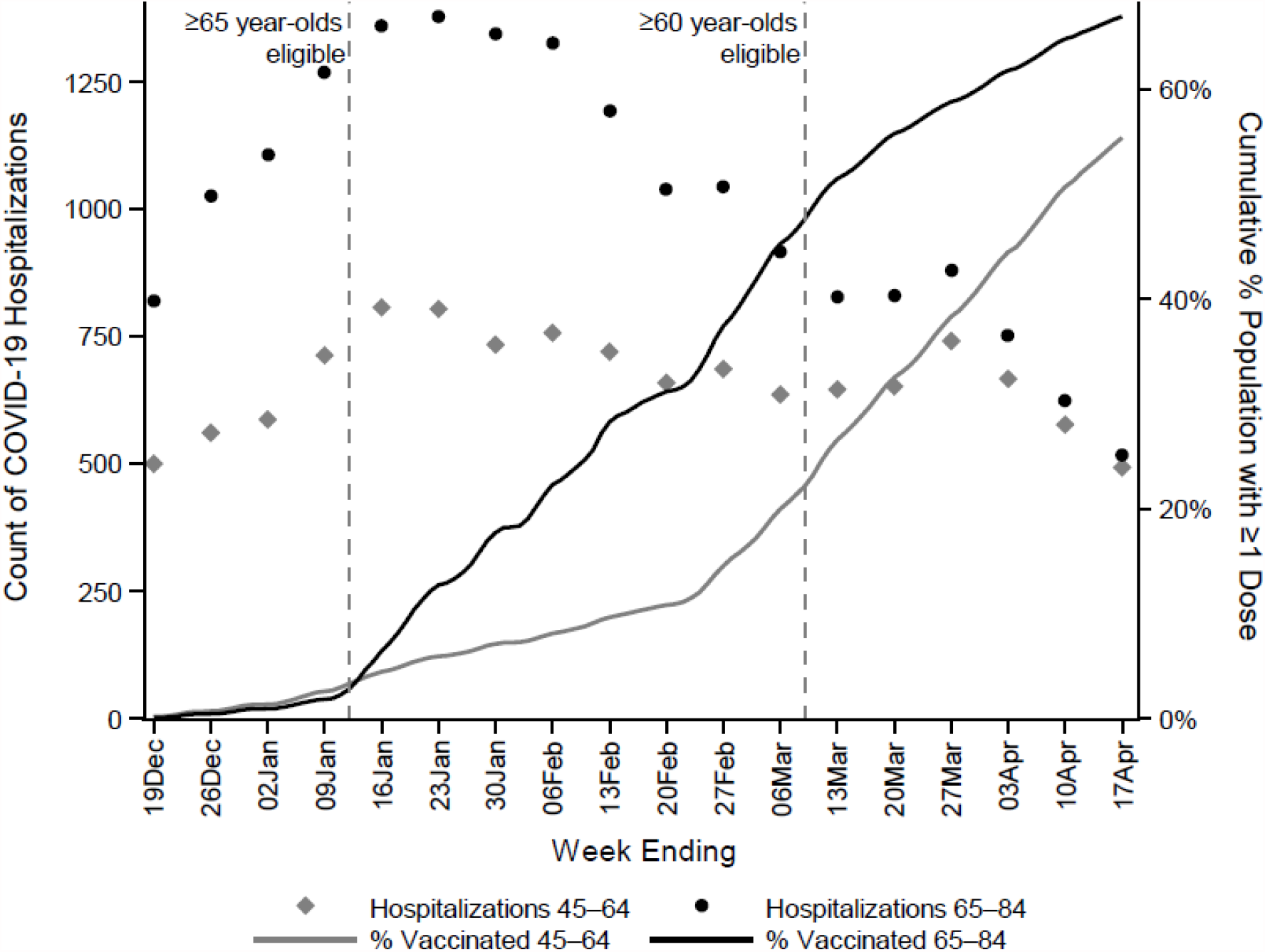
Weekly COVID-19 hospitalizations and cumulative vaccine coverage among 45–64 and 65–84-year-olds in relation to timing of age-based SARS-CoV-2 vaccine eligibility, New York City, December 13, 2020–April 17, 2021.

### 3.2 Regression discontinuity design, primary analysis

Among 2,027,014 45–64-year-old NYC residents, 5,563 COVID-19 hospitalizations occurred during the pre-implementation period and 4,977 occurred during the post-implementation period (Appendix B). Among 1,101,467 65–84-year-old NYC residents, the number of hospitalizations during pre- and post-implementation periods were 7,557 and 4,780, respectively. During the pre-implementation period, hospitalization rates increased with increasing age in years (Figure 2).

**Figure 2.**
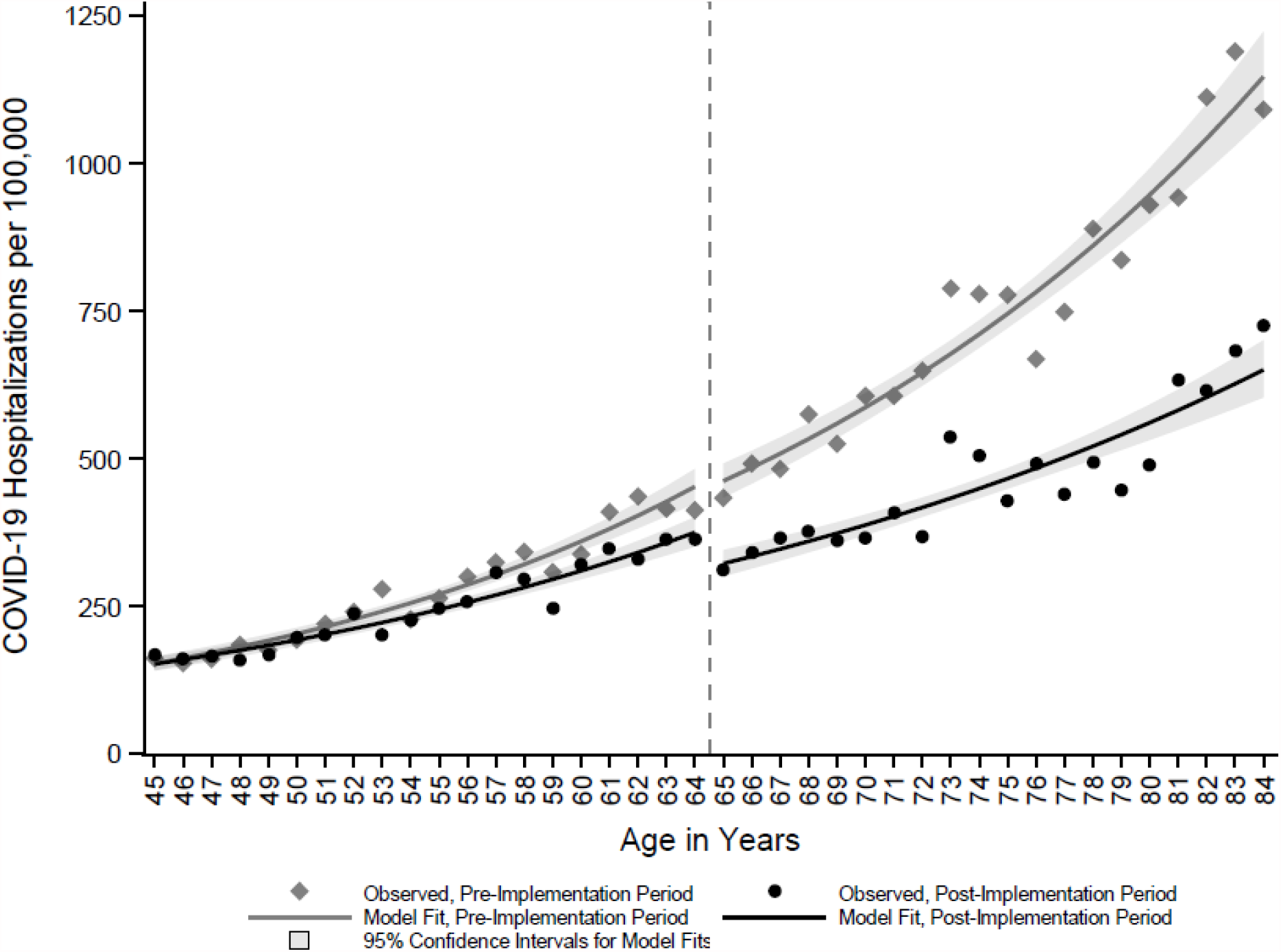
COVID-19 hospitalization rates among New York City residents by year of age during 8-week pre- (December 20, 2020– Feb 13, 2021) and post- (February 21–April 17, 2021) implementation periods for the SARS-CoV-2 vaccine program.

The hospitalization rate in the post-implementation period was lower across all ages when compared with the pre-implementation period, likely due to the waning epidemic. Even so, there was a significant negative intercept shift in the hospitalization rate of 65–84-year-olds in the post-implementation period, and 65- and 66-year-olds just above the age threshold for eligibility had lower hospitalization rates than 63- and 64-year-olds just below the threshold (Figure 2). Hospitalization rates among 65–84-year-olds during the post-implementation period decreased compared with the pre-implementation period (RR 0.85, 95% CI: 0.74–0.97, *P* = 0.02), controlling for the epidemic trend among 45–64-year-olds, a group without concurrent age-based vaccine eligibility (Table). As expected, the hospitalization rate increased with age in both pre- and post-implementation periods. The 3-way interaction between vaccine program implementation, age-based eligibility, and age in years was null, indicating that the trajectory of the hospitalization rate with increasing age did not differ before and after program implementation among 65–84-year-olds.

**Table.**
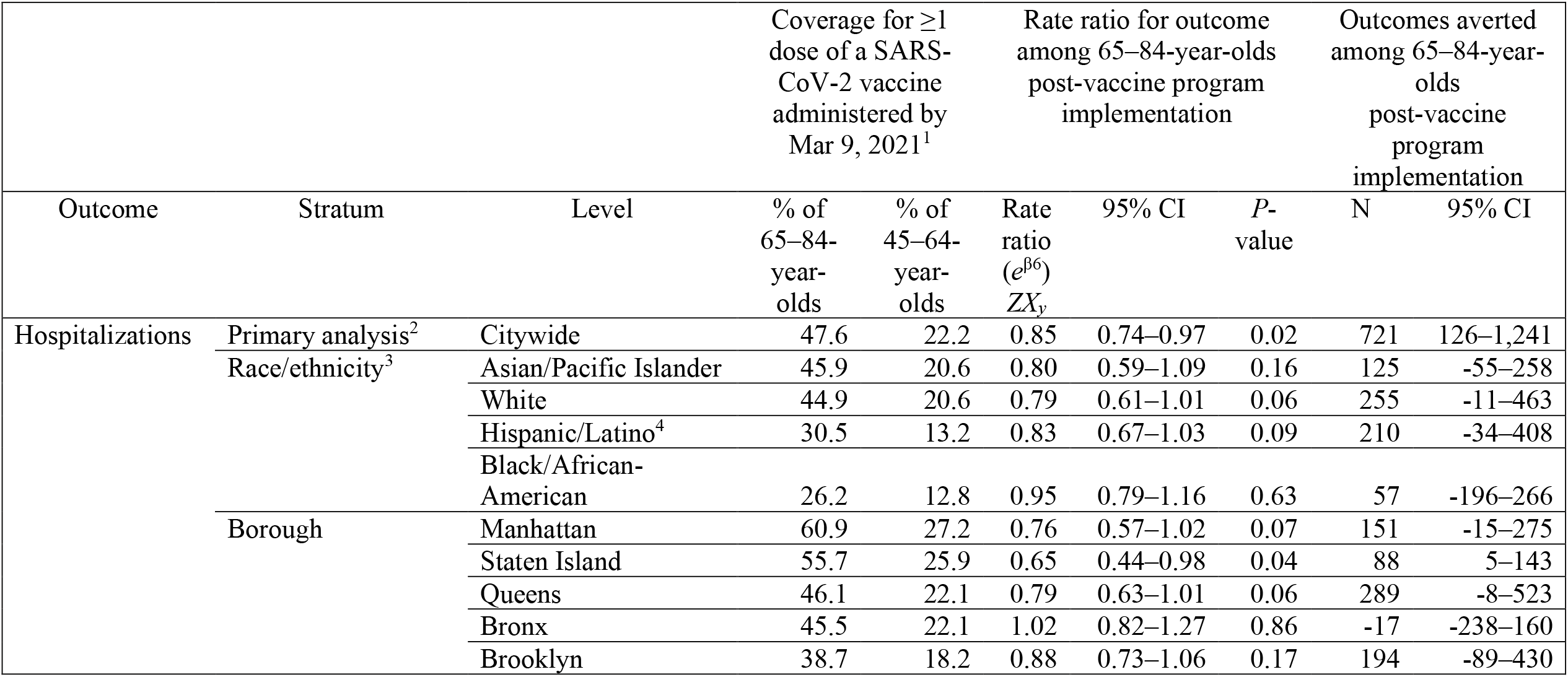
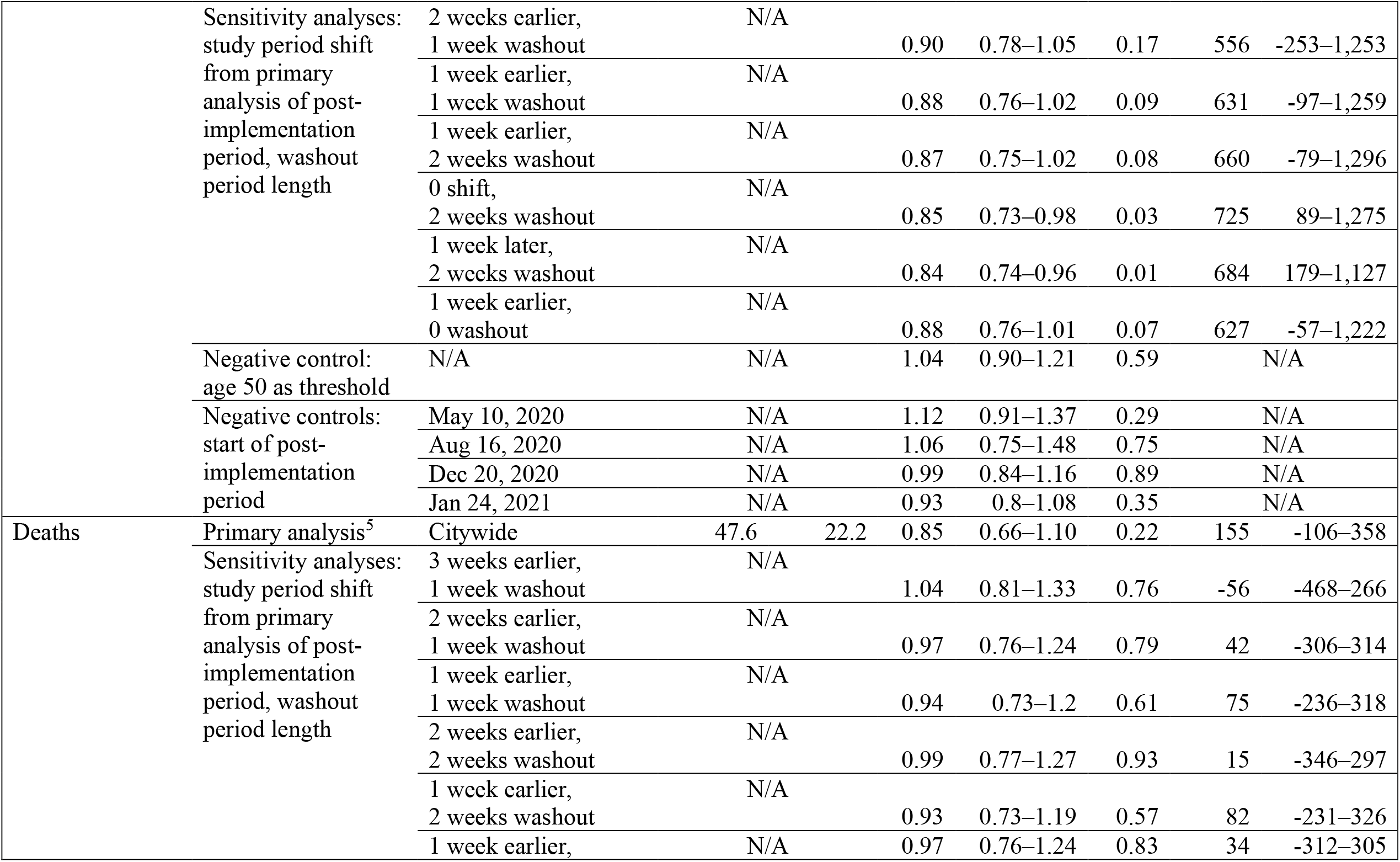
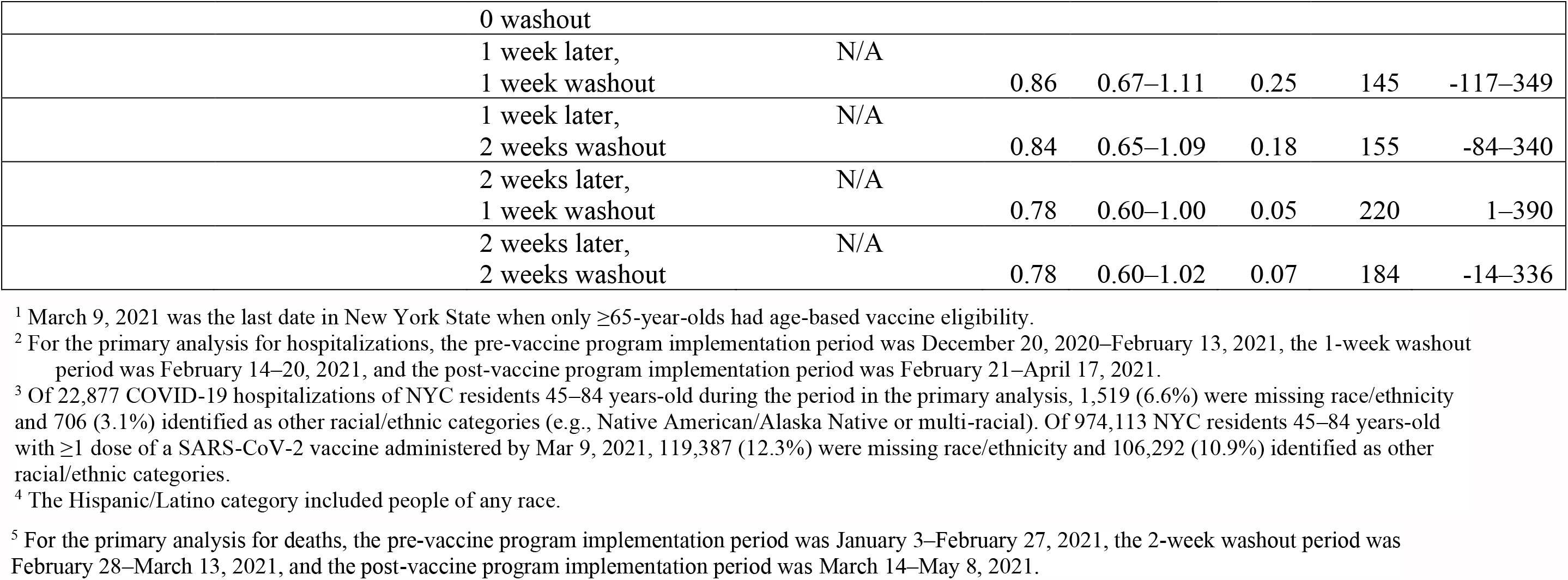
Reductions in COVID-19 hospitalizations and deaths among 65–84-year-old New York City community residents following age-based SARS-CoV-2 vaccine eligibility.

Thus, there was a 15.1% (95% CI: 2.6%–26.0%) reduction in the hospitalization rate among 65–84-year-olds during the post-implementation period compared with the pre-implementation period and accounting for the overall decrease in hospitalizations in the post-implementation period. This translates to an estimated 721 (95% CI: 126–1,241) hospitalizations averted during the 8-week post-implementation period, which are causally attributable to the vaccination program as the only intervention to reduce COVID-19 illness implemented at the ≥65-year-old threshold during this period.

In stratified analyses (Table), boroughs where residents had higher vaccination coverage generally had greater reductions in the hospitalization rate ratio point estimate, although only the citywide and Staten Island estimates were statistically significant at α = 0.05. The Bronx was an outlier, with no change in the hospitalization rate ratio (RR 1.02, 95% CI: 0.82–1.27). Black/African American individuals had the lowest vaccination coverage as of March 9 and experienced the least reduction in hospitalization rates, although differences across race/ethnic groups were not statistically significant.

Citywide during the post-implementation period, the decrease in death rates among 65– 84-year-olds was of a similar magnitude as the decrease in hospitalization rates, but deaths were sparser so uncertainty around this estimate was wide (RR 0.85, 95% CI: 0.66–1.10, *P* = 0.22) (Table).

### 3.3 Sensitivity analyses and negative controls

Findings were generally robust to program implementation period definitions. For hospitalizations, shifting the period and imposing washout periods of different lengths had minor effects on the rate ratio point estimates but influenced whether results were statistically significant. The effect for death rates was closer to the null in sensitivity analyses using earlier period definitions and further from the null using later definitions. The strongest reduction in death rates was observed when shifting the implementation period 2 weeks later and with a 1-week washout period (RR 0.78, 95% CI: 0.60–0.999, *P* = 0.05). As expected, negative controls using a false age threshold or periods prior to vaccine availability yielded no reductions in COVID-19 hospitalization rates among the older age group (Table).

## 4. Discussion

The SARS-CoV-2 vaccination program in NYC reduced the COVID-19 hospitalization rate among the initially age-eligible population by approximately 15% and was responsible for approximately 721 averted hospitalizations during the 8 weeks following program implementation. These are remarkable reductions, especially with low vaccination coverage among 65–84-year-olds during the first 8 weeks of eligibility, when demand generally exceeded supply and the cumulative percentage having received at least the first dose increased from 6.5% to only 47.6%. Our findings were robust to sensitivity analyses and negative controls.

With a regression discontinuity design, the main threat to validity would be other interventions or events to reduce COVID-19 hospitalizations that were implemented according to the same ≥65-year-old threshold and at the same time. We are unaware of any other such interventions or events, so confounding is unlikely, supporting a causal interpretation of the results [31]. The vaccination program in NYC began when the epidemic was already waning. As a point of comparison, in nearby Massachusetts, age-based eligibility began for ≥75-year-olds on February 1, 2021 [32], 1 month after the second wave of COVID-19 hospitalizations peaked in early January [33]. A strength of the regression discontinuity design (as opposed to, for example, an interrupted time series design) is avoiding the misattribution of reduced hospitalizations to vaccination as opposed to epidemic trends. Had we used time as the continuous running variable, the epidemic peak in the pre-implementation period would have complicated efforts to disentangle the vaccination program from other secular trends. While there was no discontinuity in hospitalizations over time (Fig. 1), there was a sharp discontinuity in hospitalizations by age at the vaccine eligibility threshold of 65-years-old (Fig. 2).

In stratified analyses, we observed suggestive but not statistically significant associations between increasing vaccination coverage and stronger reductions in hospitalization rate ratios. Residents of the Bronx and Brooklyn, as well as Black/African-American NYC residents, had lower vaccination coverage and appeared to experience the least reduction in COVID-19 hospitalization rates. Equitable vaccine access is urgently needed to reduce pronounced inequities in COVID-19 outcomes [34].

## 5. Limitations

The primary limitation is that our estimates of reduced COVID-19 hospitalizations in NYC attributable to the SARS-CoV-2 vaccination program are likely underestimates. The regression discontinuity design in this setting was “fuzzy” and akin to a randomized trial with imperfect adherence [35]. Vaccination coverage did not increase instantaneously following the January 12, 2021 eligibility date. Many individuals above the ≥65 years age eligibility threshold were unvaccinated, while many essential workers <65 years-old became eligible concurrently [10] and were vaccinated. A small proportion of persons who were 64 years-old as of January 12, 2021 and vaccine-ineligible would have turned 65 years-old and vaccine-eligible during the post-implementation period. Any vaccination coverage among <65-year-olds (although beneficial for vaccine recipients through the direct effects of vaccination and those around them through indirect effects [19]) would have diluted differences between older and younger groups and biased estimates of vaccine effects for the older group toward the null. Slightly faster convergence in vaccination rates between older and younger groups might partially explain why the Bronx was an outlier in stratified analyses, with no change in the hospitalization rate ratio; a mass vaccination site at Yankee Stadium, restricted to eligible Bronx residents, opened on February 5, 2021 [36], contributing to relatively quicker vaccine uptake in the Bronx among the younger age group. Similar mass vaccination sites were not opened in other boroughs until almost 3 weeks later on February 24 [37].

This study leveraged a brief, 8-week period during which age-based eligibility was restricted to ≥65-year-olds, which was only a limited period for observing reductions in hospitalization and death rates overall and any heterogeneity across subpopulations with different vaccination coverage. These estimates could not be updated using the regression discontinuity design as vaccination coverage further increased or as age eligibility expanded to younger persons, given lack of an appropriate comparator. Additionally, vaccine effects on asymptomatic or mild infections could not be assessed because vaccinated patients could have been less likely to seek testing, so estimates based on reported cases could have been biased.

Findings from the regression discontinuity design would have been further strengthened with a negative control outcome [30, 38], i.e., demonstrating no reduction among ≥65-year-olds post-vaccine program implementation in hospitalizations for a different cause also associated with age, such as myocardial infarctions. However, hospitalization data for COVID-19, but not for other causes, were ascertained through emergency response efforts to import and match data from supplemental systems [21]. Such an analysis could be explored in the future once comprehensive, all-payer hospitalization data for NYC residents become available from the Statewide Planning and Research Cooperative System (SPARCS) [39].

Finally, data included in this analysis were preliminary and subject to missing observations, missing values, and misclassification. Hospitalizations were incompletely ascertained via matching with external sources [21]. Additionally, a small proportion of hospitalizations classified as COVID-19-attributable might have been due to other causes (e.g., injuries) or were misclassified because DOHMH quality assurance processes might not have eliminated all patients who only had an encounter at an emergency department but were not admitted to a hospital. However, missing or misclassified hospitalizations would bias findings only if differential both by age and time, which is unlikely. Immunizations were incompletely ascertained for NYC residents who were vaccinated outside of New York State or by federal programs. Demographic data, notably for race/ethnicity, were incomplete for both hospitalizations and immunizations. Population denominators by single year of age were unavailable for small geographic areas or for disaggregated race/ethnic groups, limiting the ability to further examine inequities [40]. Furthermore, the denominators were vintage 2019 and did not account for any population changes between pre- and post-implementation periods or pandemic-related deaths and outmigration from NYC [41]; however, large-scale population changes during the brief period examined are unlikely, and the denominators yielded hospitalization rates with the expected positive association with increasing age during the pre-implementation period (Figure 2).

## 6. Conclusion

We demonstrated real-world evidence of SARS-CoV-2 vaccination effectiveness in protecting NYC residents in the community setting from severe COVID-19 illness. The regression discontinuity design is valid for causal inference and is low-cost to implement as an ecological, observational study with no requirement to ascertain individual-level vaccination status. This design could be used by other local and state health departments to demonstrate vaccination effects in their own jurisdictions to support public and provider messaging about the importance of vaccination. Such an approach would complement other methods for evaluating real-world vaccine effectiveness, including mathematical model-based approaches for estimating outcomes averted [42, 43] and test-negative designs [44].

## Data Availability

The analytic dataset for the primary analysis for hospitalizations is provided in Appendix B. Additional publicly available data on COVID-19 hospitalization and death rates and vaccination coverage are linked below.

https://www1.nyc.gov/site/doh/covid/covid-19-data.page

## Acknowledgments

The authors thank all NYC DOHMH staff serving in the Surveillance and Epidemiology Section and Vaccine Operations Center of the Incident Command System. We thank the Bureau of Vital Statistics for mortality data management, Iris Cheng for contributions to immunization data management, Dr. Jane Zucker for constructive manuscript review, and Dr. Annie Fine for contributions to conceptualization and data management oversight.

## Appendix A.

### Negative binomial regression model specification for controlled regression discontinuity analysis

*Y*_*y*_ = β_0_ + β_1_*A* + β_2_*X*_*y*_ + β_3_*AX*_*y*_ + β_4_*Z* + β_5_*ZA* + β_6_*ZX*_*y*_ + β_7_*ZX*_*y*_*A* + log(*N*) + e_t_, where: Y_*y*_: outcome, i.e., count (or rate, with use of offset term) of hospitalizations (or deaths) measured at each year of age *y*

β_0_: intercept, i.e., level of outcome among 45-year-olds prior to vaccine program implementation

*A*: age in years as of Jan. 12, 2021, continuous starting with 45-year-olds (i.e., 0 if 45 years, then sequentially numbered, i.e., 1 if 46, 2 if 47, … 39 if 84). β_1_ is the slope, i.e., trajectory of the outcome with each increasing year of age until age 65 years prior to vaccine program implementation.

X_*y*_: dummy variable representing vaccine program age-based eligibility (45–64-year-olds = 0, 65–84-year-olds = 1). β_2_ is the intercept for 65-year-olds prior to vaccine program implementation.

*AX*_*y*_: interaction term between age in years and age-based eligibility (0 if age ≤65, then sequentially numbered, i.e., 1 if 66, 2 if 67, … 19 if 84). β_3_ represents the change in slope or trajectory of the outcome for 65–84-year-olds relative to 45–64-year-olds prior to vaccine program implementation.

*Z*: dummy variable representing vaccine program implementation (pre-implementation period = 0, post-implementation period = 1). β_4_ represents the difference in the level (intercept) of the outcome among 45-year-olds after vaccine program implementation.

*ZA*: interaction term between vaccine program implementation and age (0 if pre-implementation period; else age indicator). β_5_ represents the difference in the trajectory (slope) of the outcome for 45–64-year-olds before and after program implementation.

*ZX*_*y*_: interaction term between vaccine program implementation and age-based eligibility (0 if 45–64 years-old or pre-implementation period; else 1 if 65–84y and post-implementation period). β_6_ represents the difference before and after program implementation in the outcome level for 65-year-olds, i.e., immediately following age-based eligibility. This is the key result of interest.

*ZX*_*y*_*A*: 3-way interaction term between vaccine program implementation, age-based eligibility, and age in years (0 if pre-implementation period or 45–65 years-old; else, if post-implementation period, 1 if 66, 2 if 67, … 19 if 84). β_7_ represents the difference before and after program implementation in the trajectory of the outcome with increasing age among 65–84-year-olds.

log(*N*): offset term, i.e., the log of the number of NYC residents by single year of age for 2019 [24].

## Appendix B.

### Analytic dataset for primary analysis for hospitalizations using the regression discontinuity design

| $Y_y$ | $A$ | $X_y$ | $AX_y$ | $Z$ | $ZA$ | $ZX_y$ | $ZX_yA$ | $N$ | $\log(N)$ |
| --- | --- | --- | --- | --- | --- | --- | --- | --- | --- |
| 161 | 0 | 0 | 0 | 0 | 0 | 0 | 0 | 99944 | 11.51237 |
| 167 | 0 | 0 | 0 | 1 | 0 | 0 | 0 | 99944 | 11.51237 |
| 151 | 1 | 0 | 0 | 0 | 0 | 0 | 0 | 98832 | 11.50118 |
| 158 | 1 | 0 | 0 | 1 | 1 | 0 | 0 | 98832 | 11.50118 |
| 161 | 2 | 0 | 0 | 0 | 0 | 0 | 0 | 100707 | 11.51997 |
| 166 | 2 | 0 | 0 | 1 | 2 | 0 | 0 | 100707 | 11.51997 |
| 193 | 3 | 0 | 0 | 0 | 0 | 0 | 0 | 105129 | 11.56294 |
| 166 | 3 | 0 | 0 | 1 | 3 | 0 | 0 | 105129 | 11.56294 |
| 193 | 4 | 0 | 0 | 0 | 0 | 0 | 0 | 110124 | 11.60936 |
| 183 | 4 | 0 | 0 | 1 | 4 | 0 | 0 | 110124 | 11.60936 |
| 198 | 5 | 0 | 0 | 0 | 0 | 0 | 0 | 102839 | 11.54092 |
| 203 | 5 | 0 | 0 | 1 | 5 | 0 | 0 | 102839 | 11.54092 |
| 222 | 6 | 0 | 0 | 0 | 0 | 0 | 0 | 101081 | 11.52368 |
| 202 | 6 | 0 | 0 | 1 | 6 | 0 | 0 | 101081 | 11.52368 |
| 239 | 7 | 0 | 0 | 0 | 0 | 0 | 0 | 99617 | 11.50909 |
| 237 | 7 | 0 | 0 | 1 | 7 | 0 | 0 | 99617 | 11.50909 |
| 284 | 8 | 0 | 0 | 0 | 0 | 0 | 0 | 102045 | 11.53317 |
| 204 | 8 | 0 | 0 | 1 | 8 | 0 | 0 | 102045 | 11.53317 |
| 245 | 9 | 0 | 0 | 0 | 0 | 0 | 0 | 107769 | 11.58775 |
| 242 | 9 | 0 | 0 | 1 | 9 | 0 | 0 | 107769 | 11.58775 |
| 281 | 10 | 0 | 0 | 0 | 0 | 0 | 0 | 106942 | 11.58004 |
| 264 | 10 | 0 | 0 | 1 | 10 | 0 | 0 | 106942 | 11.58004 |
| 314 | 11 | 0 | 0 | 0 | 0 | 0 | 0 | 104869 | 11.56047 |
| 270 | 11 | 0 | 0 | 1 | 11 | 0 | 0 | 104869 | 11.56047 |
| 328 | 12 | 0 | 0 | 0 | 0 | 0 | 0 | 101226 | 11.52511 |
| 310 | 12 | 0 | 0 | 1 | 12 | 0 | 0 | 101226 | 11.52511 |
| 345 | 13 | 0 | 0 | 0 | 0 | 0 | 0 | 100984 | 11.52272 |
| 298 | 13 | 0 | 0 | 1 | 13 | 0 | 0 | 100984 | 11.52272 |
| 325 | 14 | 0 | 0 | 0 | 0 | 0 | 0 | 105766 | 11.56898 |
| 259 | 14 | 0 | 0 | 1 | 14 | 0 | 0 | 105766 | 11.56898 |
| 336 | 15 | 0 | 0 | 0 | 0 | 0 | 0 | 99518 | 11.50809 |
| 318 | 15 | 0 | 0 | 1 | 15 | 0 | 0 | 99518 | 11.50809 |
| 399 | 16 | 0 | 0 | 0 | 0 | 0 | 0 | 97476 | 11.48736 |
| 339 | 16 | 0 | 0 | 1 | 16 | 0 | 0 | 97476 | 11.48736 |
| 420 | 17 | 0 | 0 | 0 | 0 | 0 | 0 | 96383 | 11.47609 |
| 317 | 17 | 0 | 0 | 1 | 17 | 0 | 0 | 96383 | 11.47609 |
| 385 | 18 | 0 | 0 | 0 | 0 | 0 | 0 | 92818 | 11.4384 |
| 336 | 18 | 0 | 0 | 1 | 18 | 0 | 0 | 92818 | 11.4384 |
| 383 | 19 | 0 | 0 | 0 | 0 | 0 | 0 | 92945 | 11.43976 |
| 338 | 19 | 0 | 0 | 1 | 19 | 0 | 0 | 92945 | 11.43976 |
| 373 | 20 | 1 | 0 | 0 | 0 | 0 | 0 | 86105 | 11.36332 |
| 268 | 20 | 1 | 0 | 1 | 20 | 1 | 0 | 86105 | 11.36332 |
| 399 | 21 | 1 | 1 | 0 | 0 | 0 | 0 | 81196 | 11.30462 |
| 276 | 21 | 1 | 1 | 1 | 21 | 1 | 1 | 81196 | 11.30462 |
| 377 | 22 | 1 | 2 | 0 | 0 | 0 | 0 | 78192 | 11.26692 |
| 286 | 22 | 1 | 2 | 1 | 22 | 1 | 2 | 78192 | 11.26692 |
| 438 | 23 | 1 | 3 | 0 | 0 | 0 | 0 | 76195 | 11.24105 |
| 287 | 23 | 1 | 3 | 1 | 23 | 1 | 3 | 76195 | 11.24105 |
| 406 | 24 | 1 | 4 | 0 | 0 | 0 | 0 | 77376 | 11.25643 |
| 279 | 24 | 1 | 4 | 1 | 24 | 1 | 4 | 77376 | 11.25643 |
| 437 | 25 | 1 | 5 | 0 | 0 | 0 | 0 | 72111 | 11.18596 |
| 264 | 25 | 1 | 5 | 1 | 25 | 1 | 5 | 72111 | 11.18596 |
| 424 | 26 | 1 | 6 | 0 | 0 | 0 | 0 | 69942 | 11.15542 |
| 286 | 26 | 1 | 6 | 1 | 26 | 1 | 6 | 69942 | 11.15542 |
| 446 | 27 | 1 | 7 | 0 | 0 | 0 | 0 | 68718 | 11.13777 |
| 253 | 27 | 1 | 7 | 1 | 27 | 1 | 7 | 68718 | 11.13777 |
| 435 | 28 | 1 | 8 | 0 | 0 | 0 | 0 | 55176 | 10.91828 |
| 296 | 28 | 1 | 8 | 1 | 28 | 1 | 8 | 55176 | 10.91828 |
| 419 | 29 | 1 | 9 | 0 | 0 | 0 | 0 | 53784 | 10.89273 |
| 271 | 29 | 1 | 9 | 1 | 29 | 1 | 9 | 53784 | 10.89273 |
| 382 | 30 | 1 | 10 | 0 | 0 | 0 | 0 | 49135 | 10.80233 |
| 210 | 30 | 1 | 10 | 1 | 30 | 1 | 10 | 49135 | 10.80233 |
| 327 | 31 | 1 | 11 | 0 | 0 | 0 | 0 | 48913 | 10.7978 |
| 240 | 31 | 1 | 11 | 1 | 31 | 1 | 11 | 48913 | 10.7978 |
| 331 | 32 | 1 | 12 | 0 | 0 | 0 | 0 | 44228 | 10.69711 |
| 194 | 32 | 1 | 12 | 1 | 32 | 1 | 12 | 44228 | 10.69711 |
| 364 | 33 | 1 | 13 | 0 | 0 | 0 | 0 | 40917 | 10.6193 |
| 202 | 33 | 1 | 13 | 1 | 33 | 1 | 13 | 40917 | 10.6193 |
| 338 | 34 | 1 | 14 | 0 | 0 | 0 | 0 | 40413 | 10.60691 |
| 180 | 34 | 1 | 14 | 1 | 34 | 1 | 14 | 40413 | 10.60691 |
| 341 | 35 | 1 | 15 | 0 | 0 | 0 | 0 | 36667 | 10.50963 |
| 179 | 35 | 1 | 15 | 1 | 35 | 1 | 15 | 36667 | 10.50963 |
| 327 | 36 | 1 | 16 | 0 | 0 | 0 | 0 | 34693 | 10.45429 |
| 220 | 36 | 1 | 16 | 1 | 36 | 1 | 16 | 34693 | 10.45429 |
| 349 | 37 | 1 | 17 | 0 | 0 | 0 | 0 | 31358 | 10.35322 |
| 193 | 37 | 1 | 17 | 1 | 37 | 1 | 17 | 31358 | 10.35322 |
| 349 | 38 | 1 | 18 | 0 | 0 | 0 | 0 | 29327 | 10.28626 |
| 200 | 38 | 1 | 18 | 1 | 38 | 1 | 18 | 29327 | 10.28626 |
| 295 | 39 | 1 | 19 | 0 | 0 | 0 | 0 | 27021 | 10.20437 |
| 196 | 39 | 1 | 19 | 1 | 39 | 1 | 19 | 27021 | 10.20437 |

